# Chronic skin ulcers, Burkina Faso: review of consultation trends and patient types treated between 2013 and 2023 in the dermatology departments of Sourô Sanou and Yalgado Ouedraogo University Hospitals

**DOI:** 10.64898/2026.04.07.26350370

**Authors:** Koné Alicia Christiana, Millogo Anselme, Tranchot-Diallo Juliette, Dabré Nourrédine Aristote Wendpanga, Diallo Boukary, Konaté Issouf, Kaboré Delwendé Samuel, Tioye Yéri Lydie, Kirakoya Madi, Ouedraogo Abdoulaye, Savadogo Madi, Bamba Sanata, Zoungrana Jacques, Kagoné Thérèse, Ouedraogo Abdoul-Salam, Andonaba Jean Baptiste, Ouedraogo Macaire, Niamba Pascal

**Affiliations:** Université Nazi Boni, Bobo-Dioulasso, Burkina Faso; Centre Hospitalier Universitaire Sourô Sanou (CHUSS), Bobo-Dioulasso, Burkina Faso; Université Catholique de l’Afrique de l’Ouest, Unité Universitaire à Bobo-Dioulasso, Burkina Faso; Centre MURAZ, Institut National de Santé Publique, Burkina Faso; CHU Yalgado Ouedraogo (CHUYO), Ouagadougou, Burkina Faso; Université Joseph Ky Zerbo, Ouagadougou, Burkina Faso; Institut de recherche en sciences de la santé (IRSS/CNRST), Ouagadougou, Burkina Faso

**Keywords:** Chronic skin ulcers, leg ulcers, Buruli ulcer, Neglected tropical diseases, Burkina Faso

## Abstract

Social stigma surrounding chronic skin Ulcer leads patients to hide their wounds or delay seeking medical care. The aim of this study was to explore the types and causes of chronic skin ulcers among patients seen in the dermatology departments of two university hospitals in Burkina Faso. This was a cross-sectional, retrospective study covering an 11-year period, from 2013 to 2023. A review of consultation records allowed for the collection of sociodemographic and clinical data from 104 patients who were seen for chronic skin ulcers over the 11-year period, averaging 9 patients per year. The patients were primarily adults (n=60) and older adults (n=21). Leg ulcers were the condition observed in most patients (n=59). Eight cases of Buruli ulcer (7.69%) were identified among the 104 patients. Five of the eight cases, or 62.50%, were aged between 0 and 19 years. Half of the eight patients resided in Ouagadougou. These results highlight low utilization of dermatology services for chronic skin ulcers. Furthermore, indigenous cases of Buruli ulcer have been identified in Burkina Faso. Consequently, our findings call for the implementation of strategies focused on addressing social perceptions of these ulcers and on the screening and management of this disease.

## Introduction

Skin ulcers are wounds that appear on the skin or mucous membranes and can become chronic [1,2]. Their global distribution varies from one region to another and depends on demographic, environmental, and socioeconomic factors [1,3]. The lower extremities remain the most common site, primarily the leg [4,5,6]. Chronic forms cause human suffering such as disabilities, stigmatization, school dropout, and inactivity among victims, with significant socioeconomic consequences [7]. The causes are varied. Skin ulcers of vascular origin (venous or arterial ulcers) are the most common [1,4]. Other causes exist, including cancer, diabetes [1, 4, 8], trauma, allergies, or certain infections [4,5, 8,9,10,11]. The incidence of chronic skin ulcers increases with age and is linked to the prevalence of lifestyle-related diseases such as obesity, diabetes, and cardiovascular diseases [12, 13].

In Burkina Faso, skin ulcers are common, but few hospital-based studies have been conducted on chronic forms to determine the frequency of consultations for this condition, the patient demographics, and the various causes. It is in this context that we conducted the present study to contribute to the documentation of the epidemiology of chronic skin ulcers among patients treated in the Dermatology departments of the CHUSS in Bobo-Dioulasso and the CHUYO in Ouagadougou.

## Materials and Methods

### Ethical Considerations

The study received approval from the Ethics Committee for Health Research (CERS) of the Ministry of Health of Burkina Faso under decision no. 2026-01-0034. The data collection forms did not include names in order to ensure patient anonymity.

### Study Design, Data Collection, and Processing

This cross-sectional study with retrospective data collection was conducted in Burkina Faso, specifically in the dermatology departments of CHUSS and CHUYO. A data collection form was developed to gather information from patients who were seen between 2013 and 2023 (11 years) and who presented with chronic skin ulcers. The main variables for which data were collected concerned sociodemographic and clinical characteristics, as well as the etiologies of the ulcers. Consultation records and patient charts served as documentary sources. The collected data were entered into “Microsoft Excel 2019.” Tables and figures relating to sociodemographic characteristics and patient typology were generated using the software “Epi Info 7, version 7.2.6, 2023.” The software “QGIS 3.40 Bratislava” (https://qgis.org/download/) was used to map cases of Buruli ulcer.

## Results

### 1. Incidence of consultations for chronic skin ulcers in dermatology departments at CHUSS and CHUYO, 2013–2023

A total of 104 patients were seen in the dermatology departments of CHUSS (n=31) in Bobo-Dioulasso and CHUYO (n=73) in Ouagadougou for skin ulcers, representing an average of nine (09) patients seen per year. The number of patients seen in 2013 was 17, followed by 15 in 2018. From 2020 to 2021, the number of patients seen in Dermatology for skin ulcers decreased from five (05) to two (02) patients (Fig. 1).

**Figure 1:**
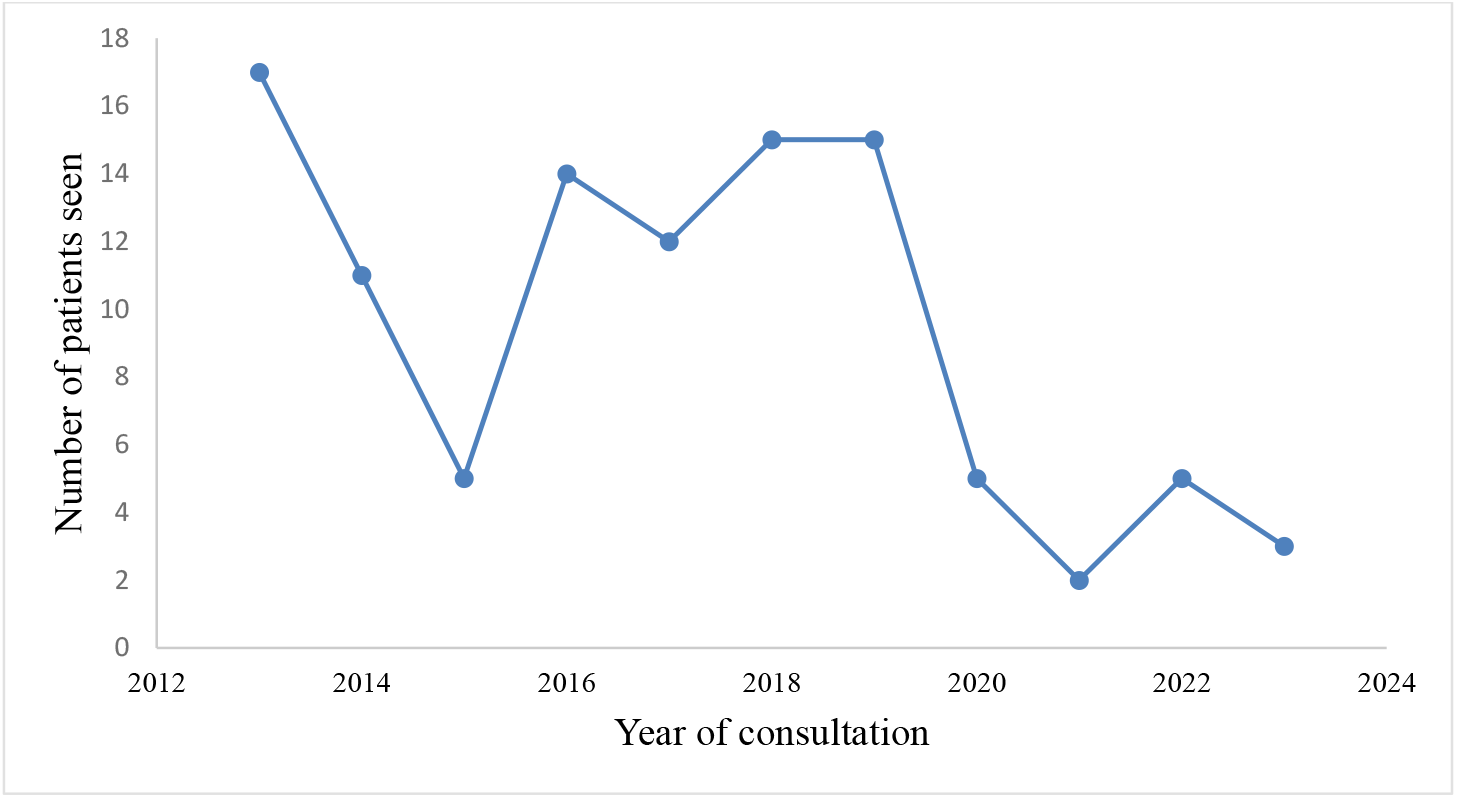
Frequency of dermatology consultations for chronic skin ulcers at CHUSS and CHUYO, 2013–2023

### 2. Patient classification by sociodemographic profile

The 104 patients were classified by age into three groups: the children and adolescents’ group, comprising patients aged 0 to 19 years; the adults’ group, comprising patients aged 20 to 59 years; and the elderly group, comprising patients aged 60 years and older [14]. There were six (06) patients in the children and adolescent’s category, and 55 in the adult’s category, representing 53.40% (55/104) of the study population. There were 21 patients aged 60 and older, representing 19.42% (21/104). Age information was missing for 22 patients (21.15%). There were 39 female subjects (37.50%), corresponding to a male-to-female ratio of 1.67 (65/39). Urban residents constituted the majority, with an overall frequency of 74.04% (77/104), and were primarily from Bobo-Dioulasso and Ouagadougou (Table I).

**Table 1.** Distribution of the 104 patients in the dermatology departments of CHUSS and CHUYO by place of residence, 2013–2023.

| Residence | Number of patients | Percentage (%) |
| --- | --- | --- |
| Urban | 77 | 74,04% |
| Rural | 16 | 15,38 % |
| Not specified | 11 | 10,58 |
| Total | 104 | 100% |

### Classification of patients based on the location of skin lesions and clinical diagnosis

Clinically, in 100 of the 104 patients, skin ulcers appeared on the skin without any particular swelling or deformation at the affected site. In three (03) patients, the ulcers were observed on nodules (ulcerated nodules), while in one (01) patient, the ulcer was associated with oedema.

The ulcers were predominantly located on the lower extremities, with 88.46% on the leg. The frequency of localization on the upper extremities was 1.92%. In 10 patients (9.62%), information regarding the location of the skin ulcers was not provided (Table 2).

**Table 2.** Distribution of the 104 patients according to lesion location.

| Location | Number of Patients | Frequency |
| --- | --- | --- |
| Leg ulcer | 92 | 88,46% |
| Arm ulcer | 2 | 1,92% |
| Not reported | 10 | 9,62% |
| Total | 104 | 100% |

Several causes were suggested in the clinical diagnosis. In 59 patients, or 56.73% of the total, a diagnosis of unspecified leg ulcer was suggested, while for 8.65% (9/104) of the patients, a diagnosis of leg ulcer of venous or arterial origin was suggested. For 7.69% of the 104 patients (8/104), a diagnosis of Buruli ulcer was established. In four patients, or 3.85% of the 104 patients, a diagnosis of venous or arterial leg ulcer or carcinoma was suggested instead. The various clinical diagnoses established are illustrated in Figure 2.

**Figure 2.**
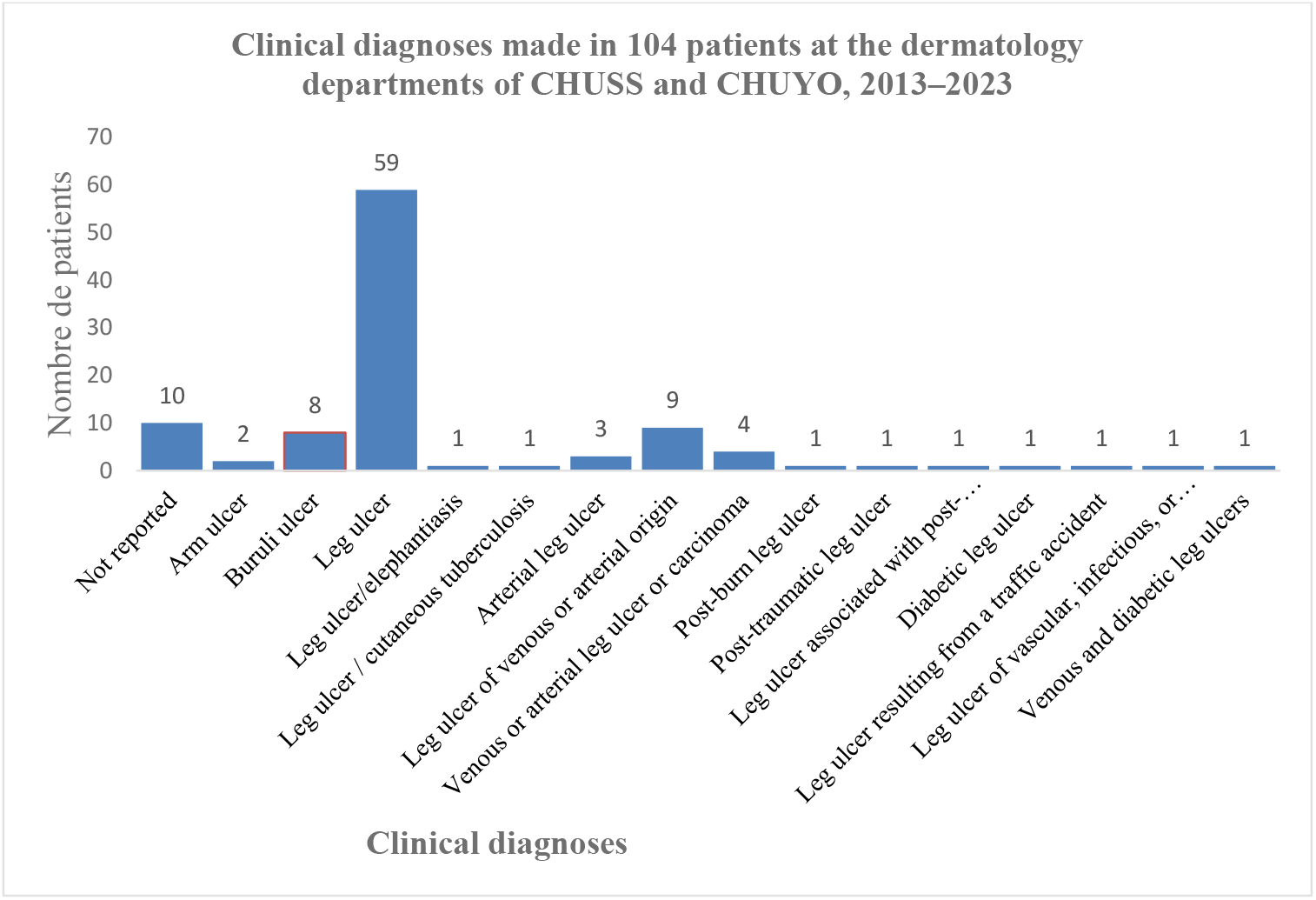
Distribution of the 104 patients according to clinical diagnoses reported in the Dermatology Departments at CHUSS and CHUYO, 2013–2023.

### 4. Typology and mapping of Buruli ulcer cases

A diagnosis of Buruli ulcer was established for eight (08) of the 104 patients, representing a frequency of 7.69% (8/104) of patients clinically diagnosed with Buruli ulcer. Six (6) of the eight (08) patients were diagnosed at CHUYO. The annual incidence of Buruli ulcer was thus estimated at approximately one (01) case per year (0.73 cases). Five of the eight patients, or 62.50%, were between the ages of 0 and 19, while the other three patients were between the ages of 20 and 59. Half of the cases were female, resulting in a sex ratio of 1. Half of the patients diagnosed with Buruli ulcer resided in Ouagadougou. The other four patients resided in Bobo-Dioulasso (1/8), Houndé (1/8), Nanoro (1/8), and Boulgou (Bagré) (1/8), as illustrated in Figure 3.

**Figure 3:**
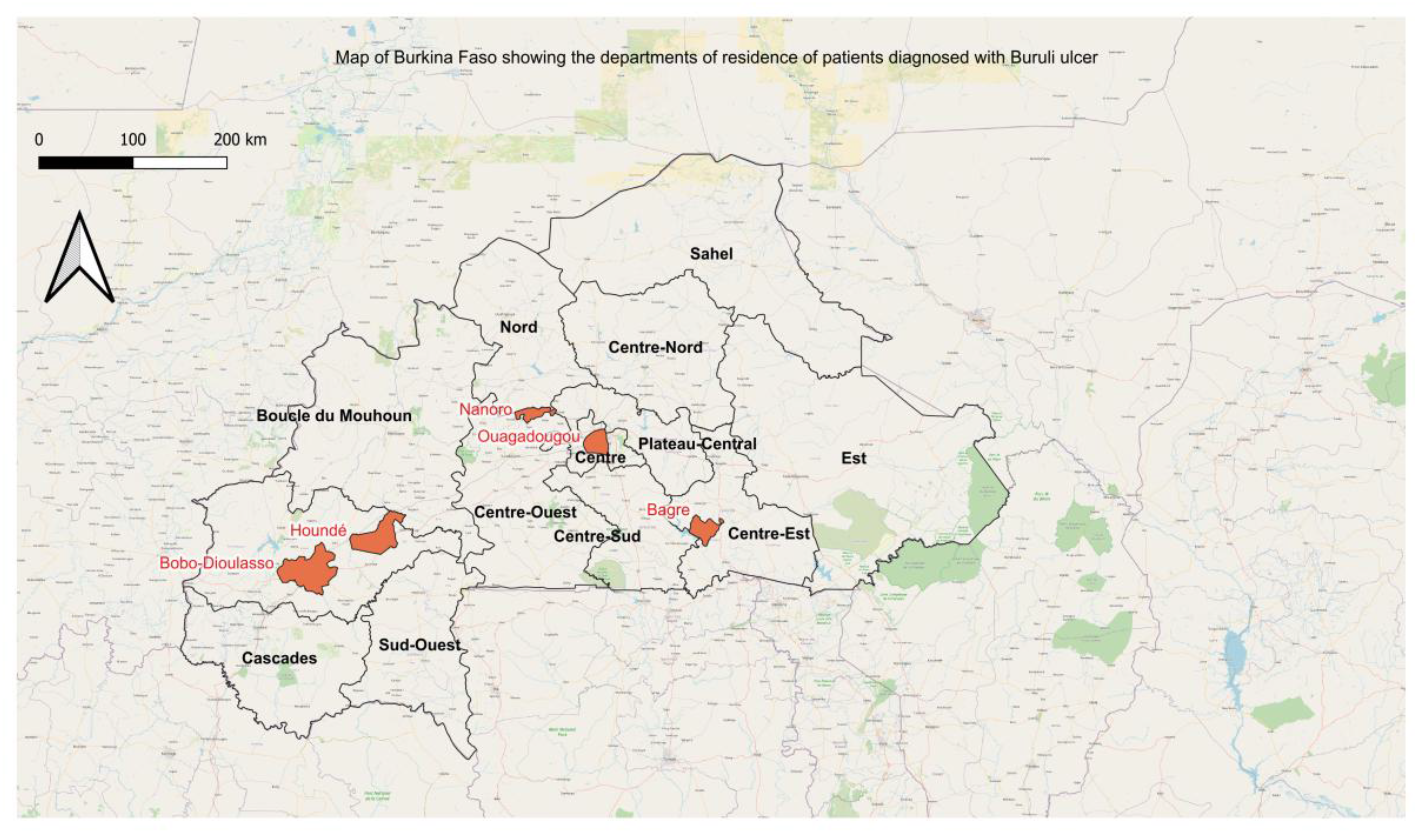
Map of Buruli ulcer cases, dermatology departments at CHUSS and CHUYO, 2013–2023. The departments of origin for patients in whom a diagnosis of Buruli ulcer was suspected are coloured red (QGIS 3.40 Bratislava (https://qgis.org/download/).

## Discussion

In our study, we observed a consultation rate of nine (09) patients per year for chronic skin ulcers during scheduled hospital visits in the Dermatology departments of CHUSS and CHUYO. These results indicate a low frequency of dermatology consultations for chronic skin ulcers and/or wounds. This could be explained by the patient care pathway in our country, where patients must first go through primary care services. Patients are referred to dermatologists at CHUSS and CHUYO only when the condition is chronic or severe [15]. Furthermore, there is the social perception of these stigmatizing conditions, whose chronic progression may lead people to believe in supernatural causes (witchcraft or transgression of social norms), resulting in a tendency among patients to hide their wounds, which may lead to low attendance at health centers [7]. The annual trend in patient visits showed a significant decline in 2020 and 2021. This situation may be due to the impact of the COVID-19 pandemic, which disrupted continuity of care for people with chronic conditions [16]. Adult patients and older adults constituted the majority of the population. Indeed, although cases of chronic skin ulcers were presented to the Pediatric department, these young patients (ages 0–19) would have been referred to and registered in the dermatology department and included in our study. The results of our study are similar to those of other studies [3,2]. Patients residing in urban areas constituted the majority. This finding could be explained by the fact that patients in rural areas are initially seen by nurses and/or general practitioners. They are referred to university hospitals only for dermatological consultations or in cases of treatment failure. The conditions diagnosed in the 104 patients were primarily chronic diseases, notably vascular ulcers and Buruli ulcer. Several authors have reported results similar to ours [2, 17, 18, 19]. The eight cases of Buruli ulcer observed in this study support the hypothesis of a probable geographic spread of this disease in West Africa, as reported by several authors [10, 11, 5].

## Conclusion

The aim of this study was to investigate the frequency of consultations in dermatology departments in Burkina Faso for chronic skin ulcers, the characteristics of the patients, and the various etiologies observed. Our results revealed a low frequency of patient consultations for chronic skin ulcers. We also report a significant decrease in consultations during the COVID-19 period, specifically between 2020 and 2021, for patients with chronic skin ulcers. The patient profile revealed that those affected were predominantly adults and males, most of whom resided in urban areas. Buruli ulcer was identified as a potential cause of chronic skin ulcers, supporting the hypothesis of a likely geographic spread of this disease in West Africa, including Burkina Faso. Our findings collectively call for public awareness campaigns to improve knowledge about chronic skin ulcers, as well as for capacity-building in medical laboratories regarding molecular diagnostic techniques for neglected tropical diseases, particularly Buruli ulcer in Burkina Faso.

## Data Availability

Our results are compiled in an Excel database in the form of tables and can be published in a supplementary table if necessary

## Acknowledgments

We would like to thank all the staff of the dermatology departments at CHUSS and CHUYO for facilitating access to patient records.

## Authors contributions

**Conceptualization:** Anselme Millogo, Juliette Tranchot-Diallo, Pascal Niamba, George Anicet Ouedraogo, Jean Baptiste Andonaba

**Data curation:** Alicia Christina Koné, Anselme Millogo, Nourrédine Aristote Wendpanga Dabré, Delwendé Samuel Kaboré

**Funding acquisition:** George Anicet Ouedraogo, Macaire Ouedraogo, Pascal Niamba, Abdoul-Salam Ouedraogo

**Investigation:** Alicia Christina Koné, Anselme Millogo

**Methodology:** Juliette Tranchot–Diallo, Issouf Konaté, Boukary Diallo, Yéri Lydie Tioye, Madi Savadogo, Thérèse Kagoné, Madi Kirakoya

**Project administration:** Juliette Tranchot–Diallo, Issouf Konaté, George Anicet Ouedraogo, Macaire Ouedraogo, Pascal Niamba, Abdoul-Salam Ouedraogo

**Validation:** Juliette Tranchot–Diallo, Issouf Konaté, Boukary Diallo,

**Supervision:** Sanata Bamba, Jacques Zoungrana, Thérèse Kagoné, Abdoul-Salam Ouedraogo, Jean Baptiste Andonaba, Macaire Ouedraogo, Pascal Niamba, George Anicet Ouedraogo

**Visualization:** Anselme Millogo, Alicia Christina Koné

**Writing original draft:** Alicia Christiana Koné, Anselme Millogo, Juliette Tranchot-Diallo

**Writing review and editing:** Alicia Christiana Koné, Anselme Millogo, Juliette Tranchot-Diallo, Nourrédine Aristote Wendpanga Dabré, Boukary Diallo, Issouf Konaté, Delwendé Samuel Kaboré, Yéri Lydie Tioye, Madi Kirakoya, Abdoulaye Ouedraogo, Madi Savadogo, Sanata Bamba, Jacques Zoungrana, Thérèse Kagoné, Abdoul-Salam Ouedraogo, Jean Baptiste Andonaba, Macaire Ouedraogo, Pascal Niamba, George Anicet Ouedraogo

